# Transcranial Ultrasound Localization Microscopy in Moya Moya patients using a clinical ultrasound system

**DOI:** 10.1101/2024.09.03.24312925

**Authors:** Louise Denis, Elena Meseguer, Augustin Gaudemer, Georges Jaklh, Sylvain Bodard, Georges Chabouh, Dominique Hervé, Eric Vicaut, Pierre Amarenco, Olivier Couture

## Abstract

**Background:** Deep brain structures are supplied by perforating arteries, these arteries are too thin to be observed with non-invasive and widely available clinical imaging methods. In Moya Moya disease, main arteries in the base of the brain progressively narrowed, and perforating arteries grow densely and tortuously to compensate the lack of blood supply in deep brain structures.

**Purpose:** The aim of this study is to evaluate the efficacy of transcranial ultrasound localization microscopy (ULM) in visualizing perforating arteries, utilizing a standard low-frame-rate ultrasound clinical scanner and contrast sequences commonly employed in hospital settings.

**Methods:** This prospective single-center study included ischemic stroke patients not related to perforating arteries, i.e. control patients, and Moya Moya disease patients (n° 2022-A02486-37). Contrast-enhanced ultrasound sequences (CEUS) were performed by an experienced neurologist and the images acquired were used to perform post-processing ULM. ULM density maps, i.e. number of microbubbles tracked per pixel, were compared with conventional 3T TOF MRI and color Doppler imaging (one-way ANOVA test). We also compared ULM density maps between the control and Moya Moya groups (two-sided parametric Student’s t-tests, or Mann-Whitney test).

**Results:** We included a group of 15 control patients and another group of 9 Moya Moya patients between March 2023 and March 2024. The patients had an average age of 45 years with 65% of them being male. Perforating arteries were captured on all subjects, with a mean diameter of 0.8 ± 0.3 mm in control patients, while it was not so far possible with TOF MRI or color Doppler (P < 0.05). Moreover, ULM enabled the differentiation between healthy subjects and those with Moya Moya disease through track mean distance (P = 0.05).

**Conclusions:** Using a low-frame-rate ultrasound scanner, CEUS and accessible post-processing tools, we have demonstrated that transcranial ULM can facilitate the visualization and characterization of perforating arteries, even in cases where they were previously undetectable using standard non-invasive imaging techniques. We speculate that with the advent of high-frame-rate 3D ULM, this technique may find widespread utility in hospitals.

**Key Results:**

- 2D low-frame rate Ultrasound Localization Microscopy (ULM) allows visualization of perforating arteries, i.e. diameter of 0.8 ± 0.3 mm.
- ULM described vessels that were not visible in conventional imaging techniques, i.e. TOF MRI and color Doppler.
- ULM reconstruction and quantification of the perforating arteries enabled the pathological group (Moya Moya) to be distinguished from control subjects.

**Summary statement:** Transcranial 2D ULM performed with a standard low frame-rate clinical ultrasound scanner enabled visualization and morphological description of perforating arteries. The study involved 24 subjects, including 9 Moya Moya patients.

## Introduction

Perforating arteries originate from the anterior choroidal artery (lenticulostriate, LSAs), the anterior cerebral artery (ACA) or the middle cerebral artery (MCA). Ranging from 0.1 to 1.2 mm in diameter (1), they are responsible of blood supply of deep brain territories (2). Perforating arteries are involved in Moya Moya disease, a rare chronic arteriopathy that arises from an occlusion, partial or complete, of the carotid artery inside the skull. Terminal carotid progressive stenosis promote the development of an additional artery network to ensure cerebral perfusion. As a result, perforating arteries find their way through various neo-anastomosis strategies (3).

Currently, arteriography of perforating arteries is used as the standard to establish the different stages of Moya Moya disease but it remains an invasive and ionizing technique (4). 7T TOF MRI has also proved effective in imaging perforating arteries, including in patients with Moya Moya symptoms (5). However, these high-field devices are not widely available in hospitals, and lower-field (1.5-3T) TOF MRIs are limited in spatial resolution for perforating arteries visualization. Transcranial ultrasound (US) is routinely used in clinical practice for non-invasive assessments. While color Doppler sequences enable visualization of the polygon of Willis and its communicating arteries, they are limited in detecting perforating arteries due to ultrasound wave diffraction (6). Contrast-enhanced ultrasound (CEUS) techniques facilitate perfusion measurements to evaluate perforating arteries’ underlying conditions but do not provide their direct visualization (7–9).

Ultrasound localization microscopy (ULM) has recently proved effective in visualizing microscopic vessels in deeper regions using US contrast agents and high-framerate sequences (10–14). In rodents, 3D ULM has been used to highlight ischemic areas (15) and transcranial 2D ULM has also been used to observe a case of cerebral aneurysm in humans (16). However, the application of transcranial 2D ULM in humans has been hindered by the requirement of a high-framerate US scanner, which is not commonly accessible in hospital settings.

The aim of this study is therefore to extend the viability of low framerate 2D ULM (17–19) for the observation and differentiation of cerebral perforating arteries between control patients without perforator involvement and patients with Moya Moya symptoms. By using conventional sequences (CEUS) and accessible tools (20) this study aims to accelerate the clinical transfer of ULM to ultrasound scanners already available in hospitals.

## Materials and methods

### Ethical approval

This clinical study was approved by the « Comité de Protection des Personnes (CPP) Ouest IV – Nantes » (Maison de la Recherche en Santé, 44000 NANTES France) on March 9, 2023. The SI identification number is: 22.04689.000108. The national identification number is: 2022-A02486-37. The ClinicalTrials.gov ID is: NCT05785598.

### Population study

This prospective, single-center, non-interventional study included 24 patients from march 2023 to march 2024. A first group of 9 patients with Moya Moya disease was selected by an expert center for this pathology (4 women and 5 men). A second group of 15 ischemic stroke patients not related to perforating arteries confirmed with MRI, i.e. control patients, was included (3 women and 12 men).

All patients underwent a clinical routine US examination, including color Doppler and CEUS sequences. Based on the patients’ temporal window and motion estimation, 21 CEUS acquisitions in total were selected for ULM post-processing, i.e. 3 Moya Moya patients were excluded based on the poor registration due to motion and poor temporal window as assessed by preliminary transcranial doppler (Figure 1, Supplementary Methods). Table 1 (Results) summarizes clinical patients characteristics, and Table 2 (Results) CEUS selection.

**Figure 1.**
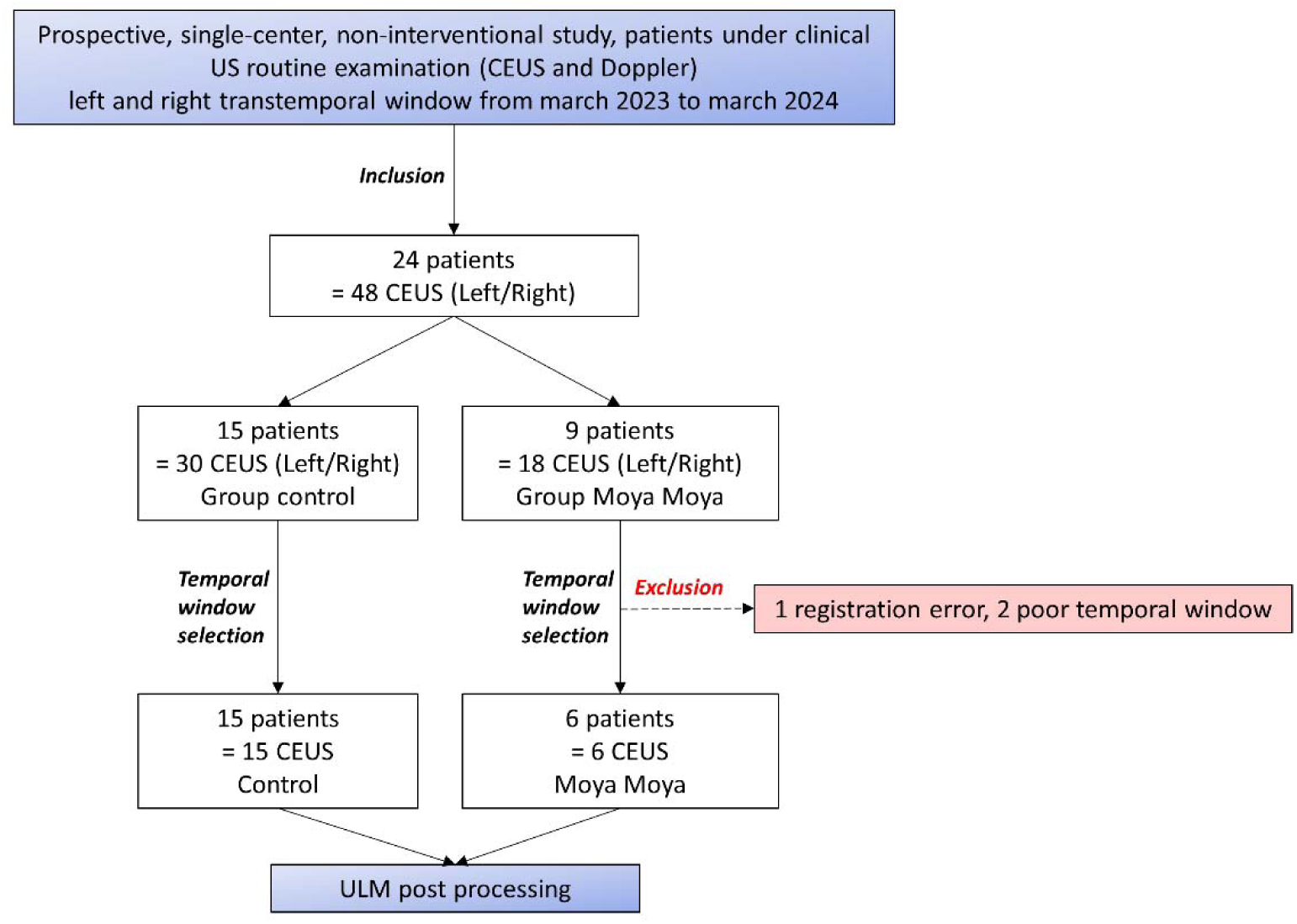
Study population. 24 patients were included, with 15 of them from a control group and 9 others with Moya Moya symptoms.

**Table 1.** Clinical characteristics of patients. FA : auricular fibrillation, FOP : permeable oval foramen, AVD : distal venous arterialization, FE : left ventricular ejection fraction

| Control patient n° | Sex | Stroke side on MRI | Thrombectomy | Thrombolysis | Stroke etiology | Included |
| --- | --- | --- | --- | --- | --- | --- |
| 1 | M | Left MCA | no | yes | cardioembolic, FOP | yes |
| 2 | M | Right MCA | no | no | atherosclerosis, stenoses carotide | yes |
| 3 | M | Right cerebellum | no | yes | atherosclerosis, occlusion AVD | yes |
| 4 | M | Left MCA | no | no | atherosclerosis, carotid stenosis | yes |
| 5 | M | Left MCA | no | no | cardioembolic FA | yes |
| 6 | M | Left MCA | no | yes | prothrombotic post heavy digestive surgery | yes |
| 7 | M | Left PCA | no | yes | cardioembolic, FE 40% apical akinesia | yes |
| 8 | M | Bilateral PICA | no | no | unknown | yes |
| 9 | M | Right PICA | no | no | unknown | yes |
| 10 | M | Left MCA | no | no | unknown | yes |
| 11 | M | Right MCA | yes | yes | cardioembolic, FA | yes |
| 12 | W | Right pontic area | no | no | unknown | yes |
| 13 | W | Left cerebellum<br>Right frontal area | no | no | cardioembolic, FOP | yes |
| 14 | M | Right MCA | no | no | unknown | yes |
| 15 | W | Left MCA | no | yes | cardioembolic, FOP | yes |
| Moya Patient n° | Sex | Moya side on arteriography | Network description |  |  | Included |
| 16 | M | Bilateral | Near occlusion termination bilateral carotid with collateral network |  |  | yes |
| 17 | W | Bilateral | Moderate Moya network |  |  | no |
| 18 | M | Left | Overdevelopment of lenticulostriate arteries in MCA left territory |  |  | yes |
| 19 | M | Right | Right carotid end stenosis and right MCA occlusion with collateral network |  |  | yes |
| 20 | W | Right | Right unilateral Moya |  |  | yes |
| 21 | M | Left | Unilateral left moya network |  |  | yes |
| 22 | M | Right | Unilateral Moya in the region of right MCA |  |  | yes |
| 23 | W | Bilateral | Bilateral Moya Moya |  |  | no |
| 24 | W | Left | Right central posterior parasagittal meningioma. Moya Moya of right carotid termination. Left ACM co-post developed. |  |  | no |

**Table 2.**
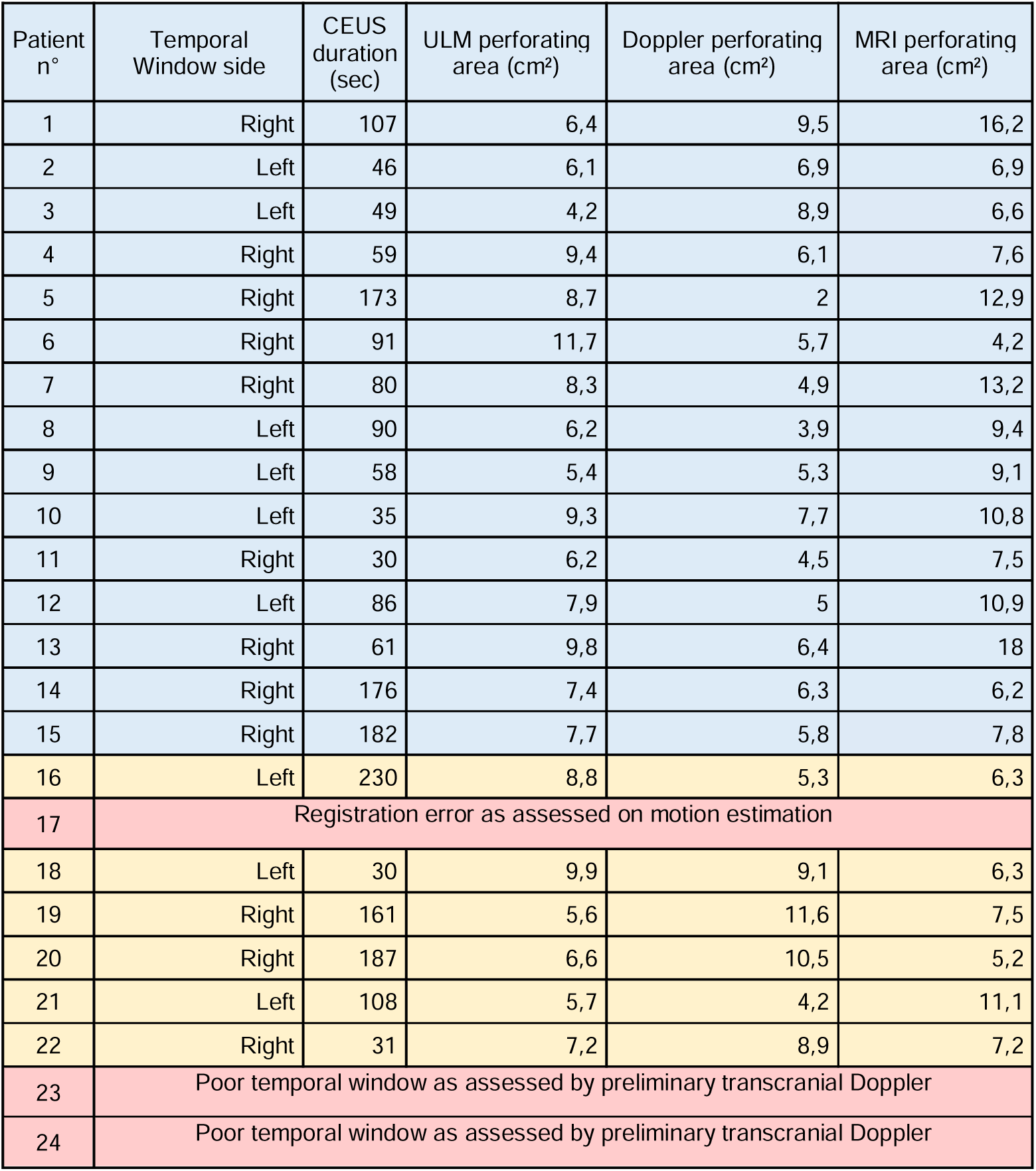
Acquisition parameters and segmented areas.

### US acquisitions

Acquisitions were performed using a clinical US scanner (Siemens Acuson S2000) and a commercial US probe (M4v1c, 2 MHz). For each patient, an initial color Doppler sequence was acquired before US contrast agent injection, to establish the quality of the acoustic window and ensure probe positioning in the MCA region (Supplementary Methods). CEUS mode was then used, as in clinical routine, alongside an intravenous injection of 2 ml bolus of Sonovue (Bracco, Italy), followed by an injection of 10cc of serum physiology. Clips from 46 to 230 seconds were saved. At the end of the bolus, a new color Doppler image was acquired, with the remaining contrast agent. The imaging depth was 9 cm, the imaging rate was 19 Hz and the clips lasted around twenty seconds each. The initial pixel resolution was 0.13 mm. The non-derated mechanical index was 0.6 and the cranial thermal index was 1.6 (in CEUS mode). Data were anonymized and then post-processed in the laboratory.

### ULM post-processing steps

CEUS clips were used to perform ULM density maps, i.e. number of microbubbles tracked per pixel, thanks to several conventional steps (11,20). First, we estimated movement and concentration using CEUS clips. Upon these measurements, we selected optimal CEUS clips, i.e. with no movement and distinguishable microbubbles (see Supplementary Materials, SF1). A filtering step was then performed by applying a time gain compensation for signal attenuation at depth. A binary perfusion mask was also applied to reduce the probability of noise localization (Supplementary materials, SF2).

Microbubbles were then localized by Gaussian correlation, i.e. normalized cross-correlation with a PSF (Point Spread Function) of two different sizes. The point spread function (PSF) was simulated at the center of a fixed-edge kernel and we adapted the edge of the kernel and the standard deviation σ of the PSF to localize both concentrated and isolated microbubbles. For concentrated microbubbles, the PSF was set to an empiric value of 10 pixels in edge length hence two microbubbles are separated by a distance of 2λ (20,21). Note that pixel size was λ/5. For isolated microbubbles, the PSF was set to 30 edge pixels, i.e. a spacing of 5λ between each microbubble, to include more isolated microbubbles. We also adjusted the standard deviation (σ) of these two PSFs to accommodate the widening of the PSF at depth, expanding from λ at the surface to 3λ at depth. Further details regarding the impact of PSF size and σ can be found in the Supplementary Materials (SF3, SF4). All the microbubbles localized were then merged and tracked using the Hungarian algorithm with a distance between microbubbles from 0.4λ to 0.6λ and a minimum duration of 0.1 sec (22,23), with a gap of 0.05 sec allowed. The track accumulation was projected onto a final grid, i.e. a pixel size of λ/5.

A comprehensive overview of all the ULM steps is provided in Figure 2.

**Figure 2.**
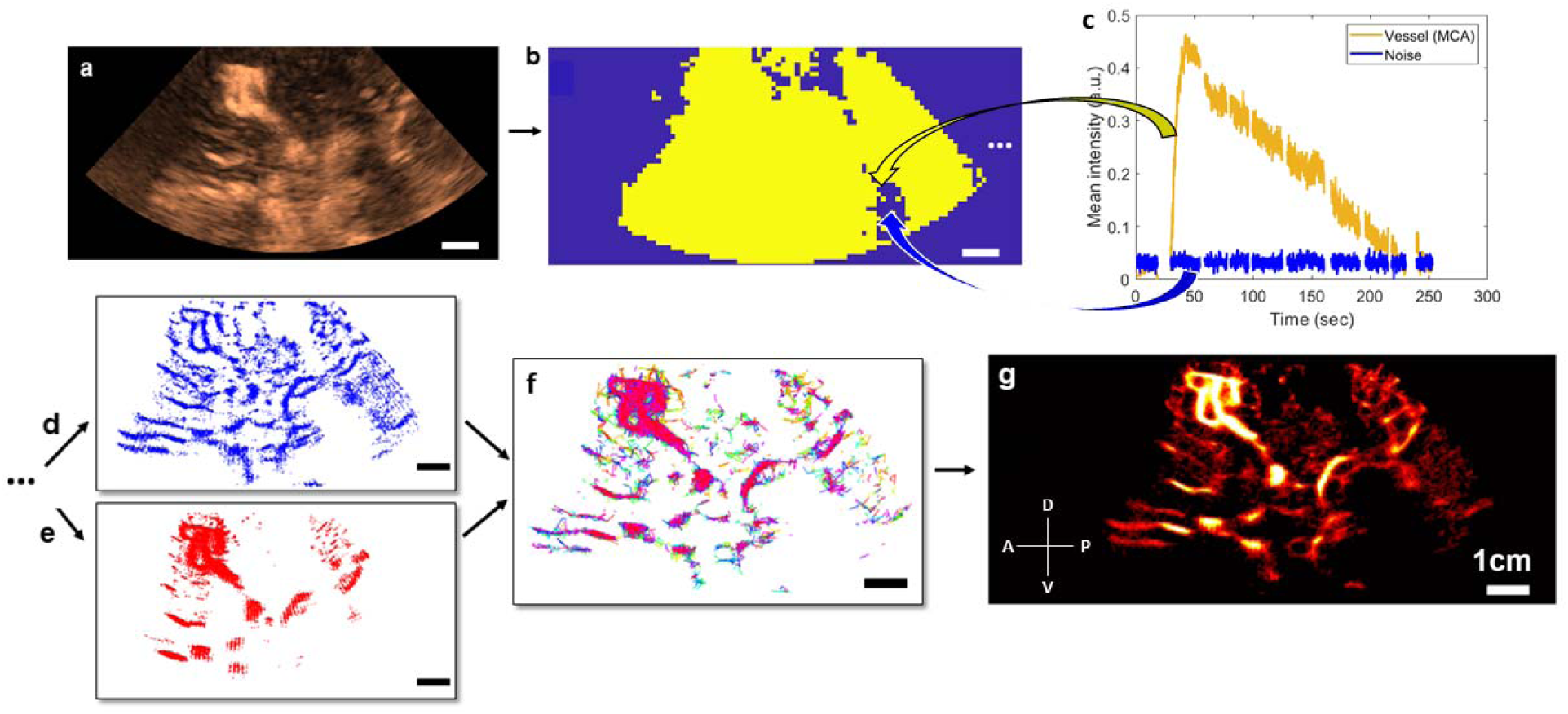
Steps of 2D transcranial ULM, example on patient n°1. a) Acquisition of clips, sorting of images with high noise level or large motion (SF1) and adjusting the TGC settings, b) Estimation of the binary perfusion mask (SF2), with typical MCA and noise patterns highlighted in c), d) Localization of concentrated MB by 10-pixel Gaussian correlation, e) Localization of isolated MB by 30-pixel Gaussian correlation, f) Tracking step with merged localization of MB, g) Accumulation of the tracks and reconstruction of a density map. Same scale everywhere: 1 cm.

### Image analysis

To estimate vessel diameters on the ULM density map, cross-sections of the vessels of interest were manually segmented. Intensity was plotted as a function of spatial distribution along the section, followed by the measurement of Full Width at Half Maximum (FWHM) (11,16).

Color Doppler acquisitions were taken from the same plane as the CEUS sequences. The 3T TOF MRI, with maximum intensity projected over 4 mm of thickness, was aligned on the ULM mapping by a neuroradiologist (CareStream software). The MCA region was manually segmented to compare the vessel density, i.e. the number of pixels above the image intensity median, in each technique.

A comparison of perforating arteries reconstructed by ULM density between control patients and Moya Moya pathology patients was carried out in the same MCA regions as ones previously manually segmented, i.e. for imaging techniques comparison. Thus, in the MCA areas, we could estimate the number of microbubbles localized, the number of tracks accumulated, the number of perforating arteries, i.e. more than 8 pixels connected, and the spatial repartition of the tracks, i.e. mean distance of all tracks to the mean point of these tracks.

### Statistical analysis

One-way ANOVA tests was used to compare ULM, TOF MRI, and color Doppler vessels’ density (Fig 4 and 5). We used unpaired two-sided parametric student’s t-tests with a 95% confidence level assuming a Gaussian distribution (Kolmogorov-Smirnov test) to compare the two groups, PSF size and σ (Fig 6 g-h and SF3). When the distribution was not normal, we used the Mann-Whitney test (Fig 6 e-f). All statistics were performed on GraphPad Prism 9 software.

## Results

### Patient characteristics

The average age of the 15 patients in the control group was around 47 years and 80% were male (Table 1). The right temporal window was chosen in 9/15 cases and CEUS clips ranged from 46 to 182 seconds (Table 2).

In the Moya Moya group, the mean age was 43 years, and the chosen time window of included patients was right temporal one in 3 out of 6 cases (Table 1). Patients were predominantly male (5 out of 6 cases) and CEUS clips were 31 seconds minimum and 230 seconds maximum (Table 2).

All these characteristics, summarized in tables 1 and 2, suggest that the two groups are comparable. In the Moya Moya group, perforating arteries development was bilateral in 3 out of 9 cases.

### ULM density maps allow the reconstruction of perforating arteries in the control group

Using post-processing tools on CEUS acquisitions at low frame rates, ULM density maps, could be reconstructed in the 15 control patients. In addition to allowing the observation of large vessels, such as the MCA and the polygon of Willis, we were able to observe perforating arteries in the region of the MCA (Figure 3a, zoomed in 3b).

**Figure 3.**
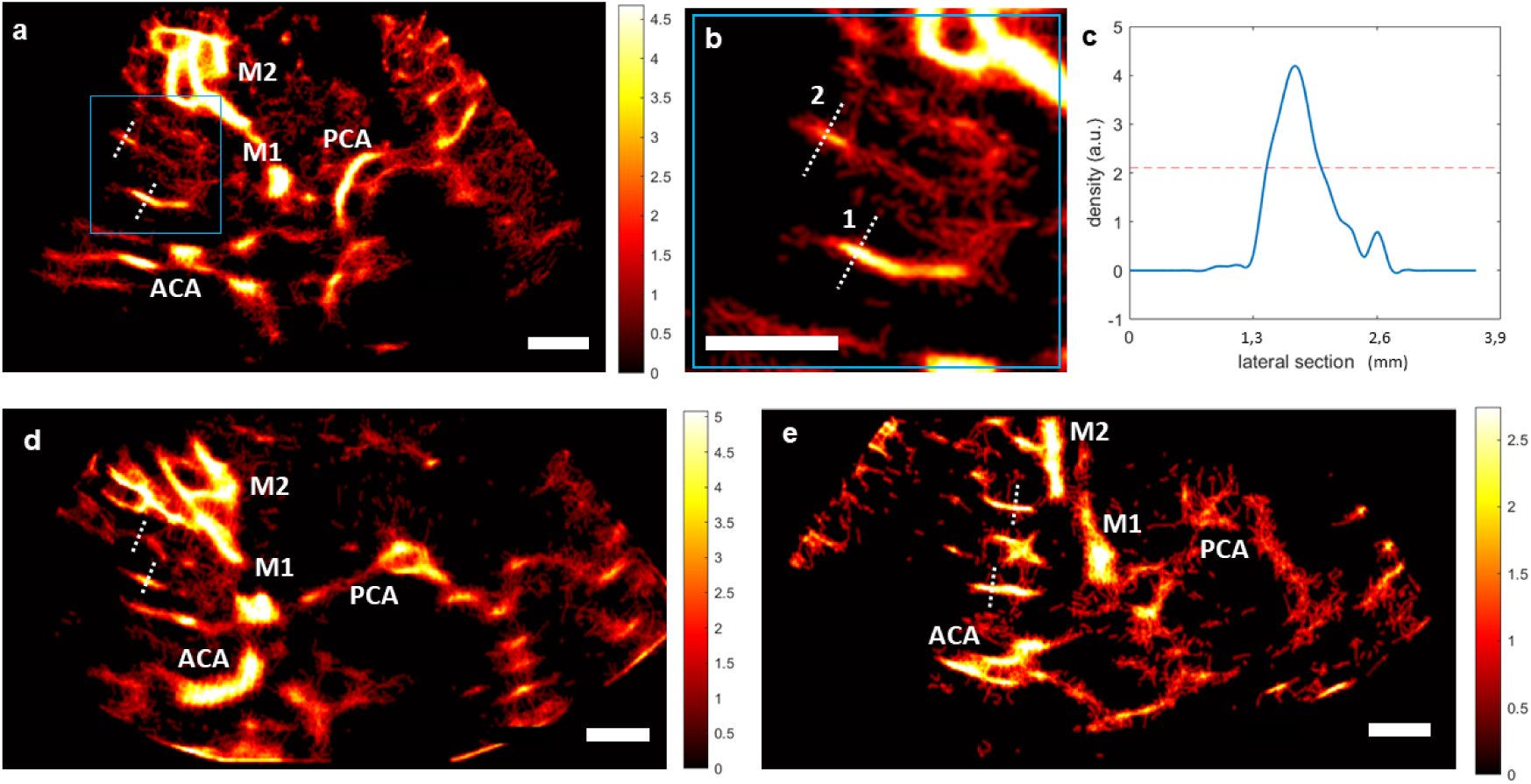
ULM density maps on control patients. a) ULM density map on patient 1, right temporal window. White dotted lines indicate segmented vessels. The blue square represents the zoomed area in b), c) Density as a function of the lateral cross-section of the first segmented vessel. The full width at half maximum (FWHM) corresponds to the vessel diameter, i.e. 0.6 mm here. d) ULM density map on patient 8, left temporal window. White dotted lines indicate segmented vessels. e) ULM density map on patient 10, left temporal window. White dotted lines indicate segmented vessels. All color bars represent the density, i.e. count of microbubbles per pixel (in the arbitrary unit, a.u.). Scale bars of 1 cm. M1: sphenoidal segment of the MCA (Middle Cerebral Artery), M2: insular segment of the MCA, ACA: Anterior Cerebral Artery, PCA: Posterior Cerebral Artery.

In each of the patients in the control group, we segmented two perforating arteries and studied the diameter of these vessels using a calculation of the full width at half maximum of the intensity (FWHM, see methods) (Figure 3 c-e). On average, the segmented perforating arteries had a width of 0.8 ± 0.2 mm in diameter, which corresponds to the size observed in the literature, i.e. from 0.1 to 1.2 mm in diameter (1).

### Comparison of ULM with MRI TOF 3T and color Doppler in the control group

The ultrasound slice plan used to perform CEUS, i.e. ULM, and color Doppler acquisitions could be manually retrieved from the 3T TOF MRI (Figures 4a, 4b). Thus, we were able to compare the MCA region of the same slice in ULM (Figure 4c), TOF MRI (Figure 4d), and color Doppler (Figure 4e).

**Figure 4.**
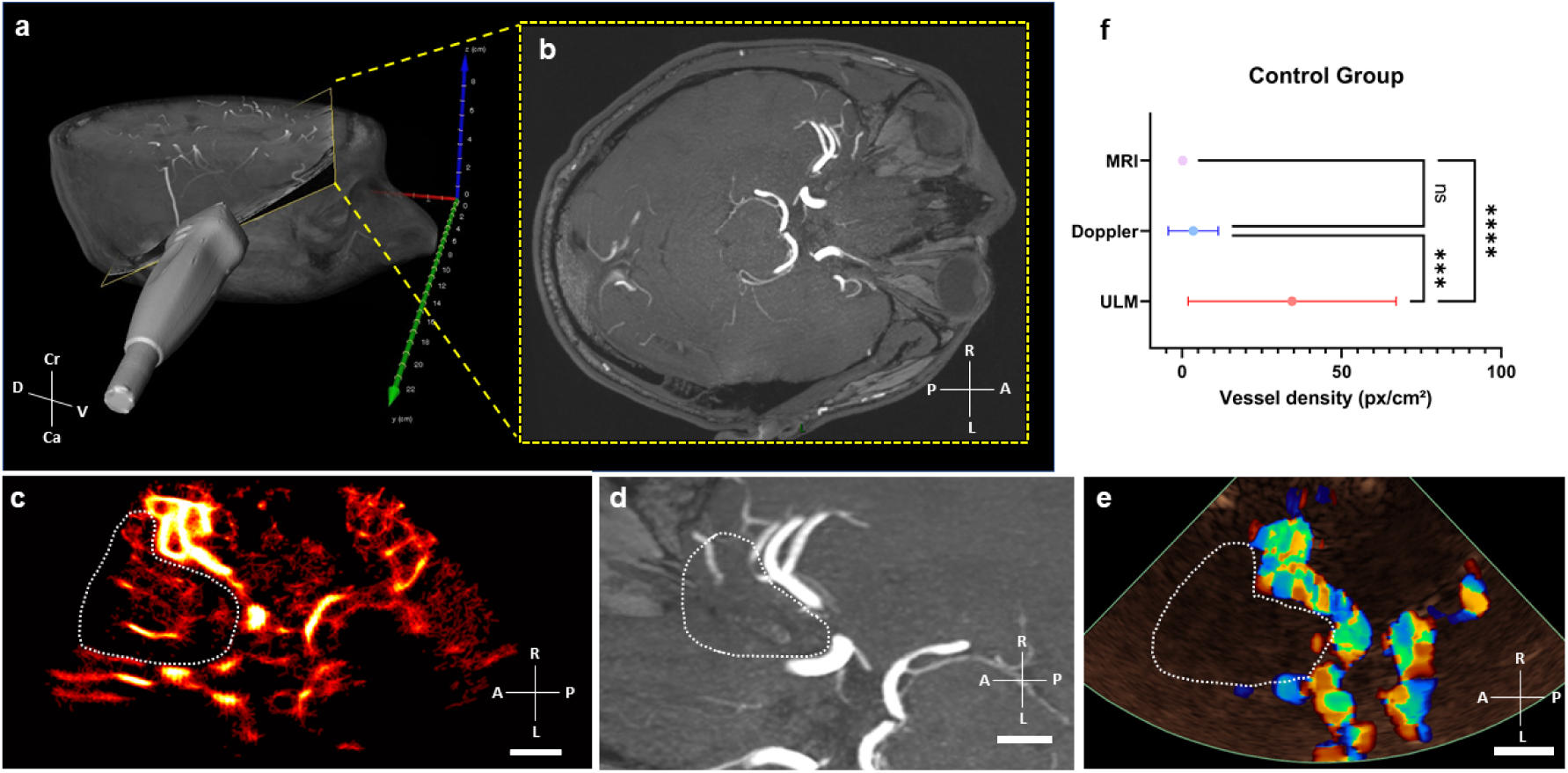
Comparison of vessel density in MRI, ULM, and color Doppler in control patients. a) Position of the phased-array ultrasound probe on the right temporal window in patient 1. Probe is superimposed on a 3D TOF MRI volume in a profile view, b) the yellow-framed slice corresponds to the registered TOF MRI slice in an axial view. c) ULM density map on the first patient (control group). White dotted lines indicate perforating arteries’ area segmentation, d) MRI TOF 3T of the same patient registered in b). White dotted lines indicate perforating arteries’ area segmentation, e) Color doppler of the same patient. White dotted lines indicate perforating arteries’ area segmentation, f) Comparison of vessel density, i.e. number of pixels superior to the median intensity of the image, between the three techniques. One-way ANOVA test (see Methods). Scale bars of 1 cm. Radiological notation, D: Dorsal, V: Ventral, Cr: Cranial, Ca: Caudal, P: Posterior, A: Anterior, R: Right, L: Left.

Overall, vessel density, calculated as the number of pixels greater than the median intensity of the image (see section Methods), is greater in ULM than in MRI and color Doppler in the MCA region in all 15 patients. Remarkably, ULM density map enables reconstruction of perforating arteries that are not visible with conventional imaging techniques used in clinical routine.

### Comparison of ULM with 3T TOF MRI and color Doppler in the Moya Moya Group

In patients in the Moya Moya group, the same 3T TOF MRI registration step was carried out with ULM and color Doppler (Figure 5 a-c). The density of reconstructed perforating arteries remained higher with ULM compared to MRI and color Doppler in 6 patients (Figure 5d), indicating the visualization of LSAs with ULM, which is not achievable with conventional imaging modalities.

**Figure 5.**
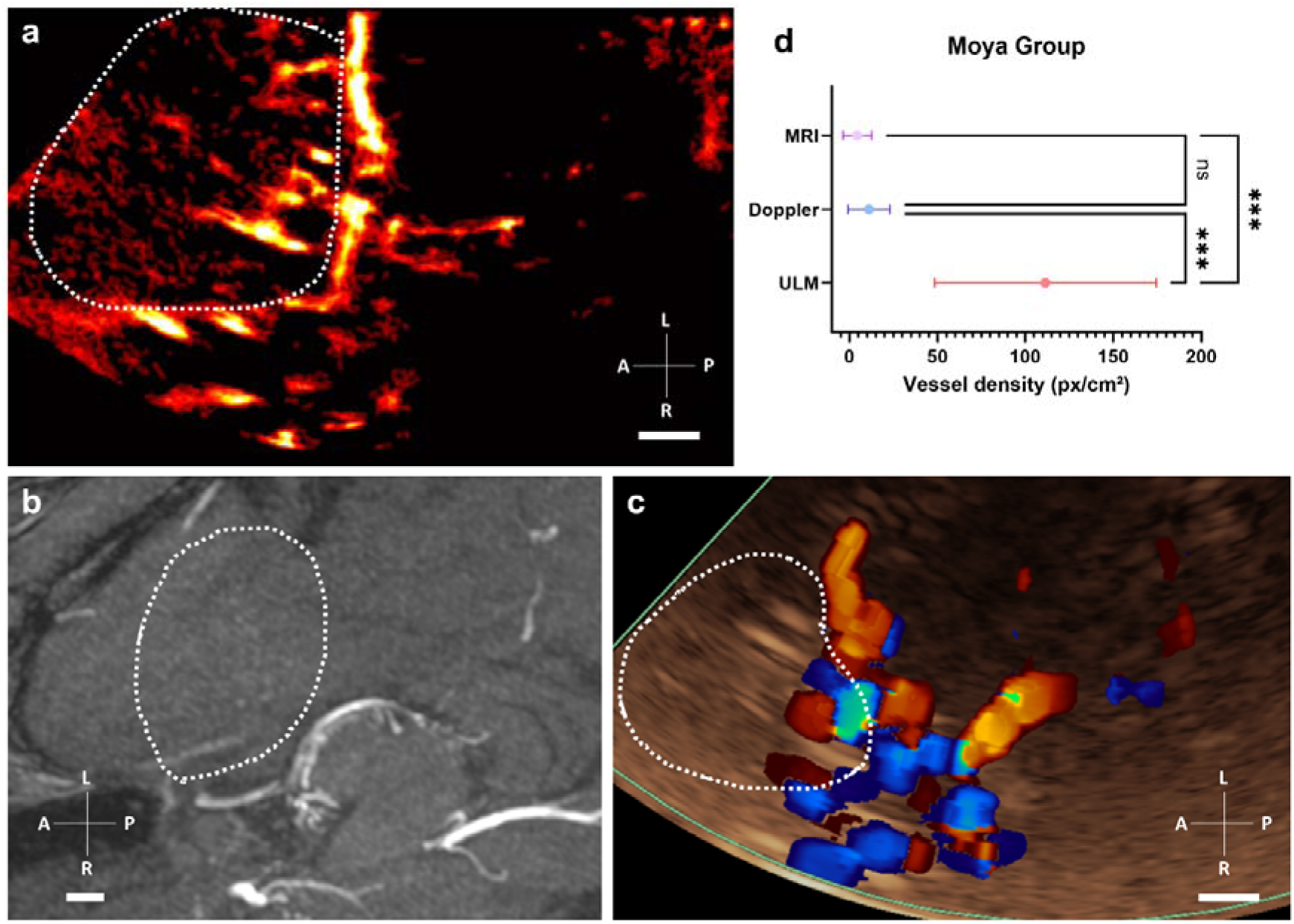
Comparison of vessel density in MRI, ULM, and color Doppler in Moya Moya patients. a) ULM density map on the patient n°16 (Moya Moya group). White dotted lines indicate perforating arteries’ area segmentation, b) MRI TOF 3T of the same patient (axial view). White dotted lines indicate perforating arteries’ area segmentation, c) Color doppler of the same patient. White dotted lines indicate perforating arteries’ area segmentation, d) Comparison of vessel density, i.e. number of pixels superior to the median intensity of the image, between the three techniques. One-way ANOVA test (see Methods). Scale bars of 1 cm. Radiological notation, P: Posterior, A: Anterior, R: Right, L: Left.

However, it is interesting to note that the mean intensity of color Doppler in the MCA region is higher in patients in the Moya Moya group than in the control group. This effect has already been observed in the literature and categorized as a characteristic pattern of disease progression (6). In this case, ULM would enable us to observe the neoangiogenic structures underlying the phase shift observed on color Doppler.

### ULM comparison between Moya Moya and the control group

The same regions segmented for comparison of imaging methods, i.e. TOF MRI, ULM, and color Doppler, were reused to compare vessels reconstructed by ULM in the control group (Figures 6 a,b) and in the group of patients with Moya Moya symptoms (Figures 6 c,d).

**Figure 6.**
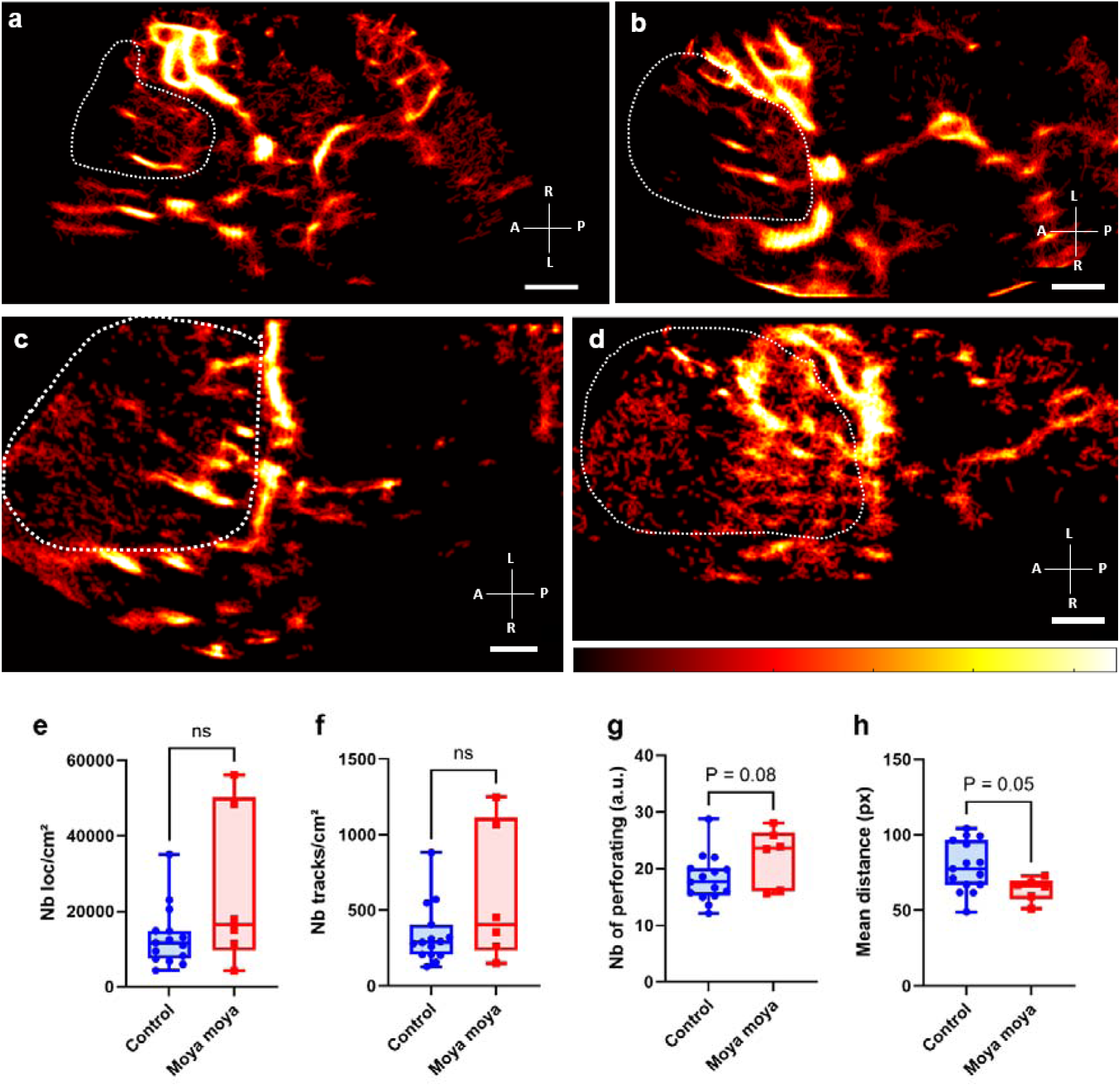
Comparison of ULM density maps between control and Moya Moya patients. a) ULM density map of control patient n°1 right temporal window, colormap from 0 to 4.5 (a.u.), b) ULM density map of control patients n°8 left temporal window, colormap from 0 to 5 (a.u.), c) ULM density map of Moya Moya patient n°16 left temporal window, colormap from 0 to 3.5 (a.u.), d) ULM density map of Moya Moya patient n°18 left temporal window, colormap from 0 to 2.5 (a.u.). e) Number of localizations per cm² inside perforating arteries’ region between Moya Moya and control patients, f) Number of tracks per cm² inside perforating arteries’ region between Moya Moya and control patients, g) Number of at least 8 pixels connected objects (a.u.), i.e. perforating arteries, inside perforating arteries’ region between Moya Moya and control patients. h) Mean distance between tracks (px) inside perforating arteries’ region between Moya Moya and control patients. Same color bar for all patients. White dotted lines indicate perforating arteries’ area segmentation. Radiological notation, P: Posterior, A: Anterior, R: Right, L: Left. Student t-tests were performed to compare the two groups (see Methods)

Overall, the number of normalized tracks per cm² and density of microbubble localizations per cm² showed no significant differences between Moya Moya patients and control patients (Figure 6e-f). Number of perforating arteries, showed no significant differences between the two groups either (P < 0.1, Figure 6g). Finally, the spatial distribution of these vessels, calculated here as the mean distance of the tracks from the mean point of all the tracks, was smaller in Moya Moya patients then in control ones (P = 0.05, Figure 6h). Vessels were therefore more numerous and populated in Moya Moya patients than in control patients.

In Moya Moya patients, we were also able to see these dense, tortuous vessels directly on some arteriography, i.e. the gold standard. Although the 2D slice limits comparison, the supply network could be found on the Moya Moya side (patient 4 in Supplementary Figure SF5 and patient 7 in Supplementary Figure SF6), and not on the control side (patient 7 Supplementary Figure 6).

## Discussion

### Summary of the entire study

In this study, we were able to demonstrate that 2D transcranial ULM density maps could be performed with conventional low frame-rate clinical ultrasound scanners. 2D transcranial ULM enabled a reconstruction of 0.8 ± 0.3 mm diameter perforating arteries in the MCA region, perforating arteries that were not visible with other conventional imaging methods, i.e. TOF MRI and color Doppler. In addition, we were able to characterize the complex vascular network of patients with Moya Moya pathology. While color Doppler highlighted pattern differences in these subjects, ULM revealed the morphological differences underlying these phase distinctions.

### Implications

The results of this study are promising regarding the capability of 2D ULM to detect fine cerebral arteries, even with the low frame rate of the acquisitions. Indeed, in addition to the cerebral perfusion that can classically be calculated on CEUS mode, ULM provides information on the layout of the vasculature underlying these vascular dynamics.

Clips from standard clinical ultrasound equipment were utilized in this study. This showcase the method’s scalability. Access to publicly available codes further enhances its applicability (20). In this way, it should be easy to explore other organs using the same algorithms coupled with CEUS mode (17).

### Limitations

One of the major limitations of this technique, and more generally on all transcranial ultrasound techniques, is that it can only be applied to patients with an appropriate acoustic window and we had to exclude 2 patients for this reason. In our study, the average age was quite low in both groups, mean of 47 years in the control group and 43 years in the Moya Moya group. Among them, 65% of the patients included were men. Nevertheless, it should be noted that, in the literature, around 20% of patients (male/female combined) do not have a sufficient acoustic window for transcranial ultrasound (24).

Another limitation comes from the fact that ULM needs microbubble tracking accumulation at the same position for several minutes, i.e. from 30 seconds to 4 minutes. As the probe was hand-held, drift was inevitable. The current solution was to remove these clips from the analyses, which reduces the amount of usable data and therefore the number of reconstructed vessels. The ULM algorithm also requires individual microbubbles visible in the clips. However, in the case of acquisition in the MCA region, this criterion cannot be met in both large arteries and perforators. Although this limitation has been partially overcome thanks to the selection of upstream clips and the localization of microbubbles using an isolability criterion, i.e. spaced by 5 λ (Supplementary Materials, SF3, SF4), it remains one of ULM’s main constraints. Another major limitation of ULM lies in the adaptation of parameters for each patient, i.e. tracks max linking distance and their min length. Depending on the visualized phenomena, ULM tracking parameters must be adapted to follow correctly the microbubbles and these differences can cause bias in the final analyses.

Furthermore, density measurements and the analysis of various vessel sizes rely on manually segmented masks, introducing subjective bias. Likewise, registration between TOF MRI and 2D ULM is conducted by a single radiologist, potentially impacting accuracy.

### Perspectives

In the future, high frame-rate ultrasound remains the technique to prioritize. Indeed, low frame rate imaging as used here has limited us to density reconstruction. Although we were able to distinguish the pathological group from that of control patients, our analyses and quantifications were based solely on morphological measurements of the vessels. On the other hand, ULM performed with ultrafast ultrasound scanners, i.e. several hundred or even thousands of images per second, could enable measurements of vessel velocities, or even pulsatility (25), which is promising for future pathologies’ biomarkers.

Besides, volumetric imaging will be essential. 3D ULM has already proved its worth in preclinical studies on numerous organs (15,26) and remains indispensable for observing complex vascular structures that are not necessarily aligned in the 2D imaging section (18). In the case of Moya Moya, 3D would enable a fine description of vessel entanglement due to the pathology’s neoangiogenesis. Similarly, in tumors, tracking microbubbles in all spatial directions could potentially enable more precise microvascular reconstruction and even better therapeutic follow-up (27).

Our study aimed to explore the current potential of transcranial 2D ULM with low frame rate clinical ultrasound scanners and open-source tools so that all hospitals could have access to this kind of technology. In the future, high-frame rate 3D ULM could become a new reference technique for research into microvascular diseases.

## Data Availability

Patients imaging data are not available due to ethical, medicamedical, and legislative considerations toward personal information.

## Abbreviations

MB: microbubbles
US: ultrasound
ULM: ultrasound localization microscopy
CEUS: contrast-enhanced ultrasound
TOF: MRI time of flight magnetic resonance imaging
LSA: lenticulostriate arteries
MCA: middle cerebral artery
ACA: anterior cerebral artery
PCA: posterior cerebral artery

## Acknowledgments

We would like to thank the staff at Bichat Hospital (APHP Paris), and more specifically Emilie Mazars Debbah for carrying out all the Sonovue injections.

We would also like to thank the URC Lariboisiére Saint-Louis Fernand-Widal and their regulatory affairs team for setting up the clinical study.

