## Supplementary Materials for "Transcranial Ultrasound Localization Microscopy in Moya Moya patients using a clinical ultrasound system"

### Supplemental Materials

#### SUPPLEMENTARY METHODS

##### Transtemporal acoustic window estimation during US acquisitions

The quality of temporal acoustic windows was not defined using quantitative criteria. This information is derived from the clinician's subjective assessment. In some patients, transcranial color Doppler examination enabled visualization of the polygon of Willis and the MCA without contrast agent injection. These patients were considered to have a good acoustic window. Others did not, and the clinician should inject microbubbles to observe these vessels.

##### Clips selection by estimating movement and concentration during CEUS acquisitions

To estimate the concentration of microbubbles in the clips, we plotted the intensity of each clip against time. In this way, we were able to select clip numbers corresponding to usable acquisitions with single microbubbles present in the perforating arteries (Supplementary Figure 1). This estimation of microbubble concentration also enabled us to eliminate excessively noisy acquisitions, i.e. with a total image standard deviation below the noise threshold, with an empirical value of 2.5.

To estimate the motion within the probe, we calculated a power Doppler per clip, by applying a bandpass filter with a frequency cutoff of [2 15] Hz to each clip and summing all the images over time. In this way, we were able to observe the potential drift by correlating the first power Doppler (reference image) with the subsequent images (Supplementary Figure 1). Overall, a correlation below 0.9 (arbitrary threshold) represented significant movement (drift or plane change). It is essential to note that this motion estimation is intimately linked to the concentration estimation, given that we use the same CEUS images: nevertheless, this step allows us to visually assess the presence of movements.

##### Perfusion mask calculation for ULM localization step

A perfusion mask was generated to mitigate the localization of noise rather than microbubbles. Indeed, the perfusion classically observed in blood vessels has a specific pattern (7–9). Noise, on the other hand, oscillates rapidly with a low standard deviation of intensity. To create this binary perfusion mask, we concatenated all the clip images and performed spatial averaging according to a kernel arbitrarily chosen as 10 x 10 pixels. We then projected the temporal maximum intensity divided by the temporal minimum intensity in each of these kernels (Supplementary Figure 2). This projection was then normalized by the maximum image-wide value of this ratio. Then, the perfusion mask was set to 1 in kernels above the median of this normalized projection ratio, and to 0 when below it. We therefore understand that if the perfusion mask is set to 0, the gap between minimum and maximum intensity is small, which would correspond to a noisy zone. Conversely, the classic vascular pattern results in a strong maximum and a weak minimum, generating regions of the perfusion mask set to 1. This binary mask was then used during the ULM localization step (Figure 2).

##### PSF analysis for ULM localization step

The localization step is crucial in ULM, as it will largely determine which vessels can be reconstructed in the final density mapping. In the case of localization in the MCA region, microbubbles tend to be highly concentrated in the large vessels (polygons of Willis and middle cerebral artery), while they are more isolated in the perforating arteries. To localize microbubbles in these different arteries at the same time (MCA and perforating arteries), we localized microbubbles "close together" by 2 $\lambda$ (10 pixels), as well as more "isolated" microbubbles, spaced 5 λ apart (30 pixels). In the 15 control patients, we independently reconstructed the ULM density mappings by performing a single localization of isolated microbubbles (SF3 a), and of closely spaced microbubbles (SF3 b). We were thus able to quantify that the localization of microbubbles spaced 2 λ apart enhanced the MCA region, while the localization of microbubbles spaced 5 λ apart enhanced the perforator region (SF3 c,d). We therefore opted for dual localization to enhance the reconstruction of both the MCA and perforating arteries (SF3 e).

In ultrasound, the lateral resolution of a point source increases with depth according to the following relationship: $Res_{Lat} = \frac{\lambda z}{D}$, with λ the probe wavelength, i.e. 0.77 mm, z the depth and D the probe aperture, i.e. 16.3 mm. The spot of a microbubble, i.e. the PSF, close to the probe will therefore be less spread out than the spot of a microbubble at depth. To include this PSF evolution, we considered that the σ of the PSF sought during the ULM localization stage, would evolve as a function of depth, i.e. a σ of 1 λ at a depth of less than 3 λ (i.e. 2mm) up to a σ of 3 λ at depths greater than 70 λ (i.e. 5cm). In Figure SF4 we compare ULM mapping reconstructed with a PSF of 30 edge pixels (5 λ) and a σ of 7 pixels (1 λ) (SF4 a), or with a σ of 25 pixels (4 λ) (SF4 b) or with a depth-adapted σ, i.e. from 5 (1 λ) to 14 pixels (3 λ) (SF4 c). The evolution of the average intensity of these depth maps confirms that adapting the size of the σ to depth enhances both high and low perforators (SF4 d).

#### SUPPLEMENTARY FIGURES (1 to 6)

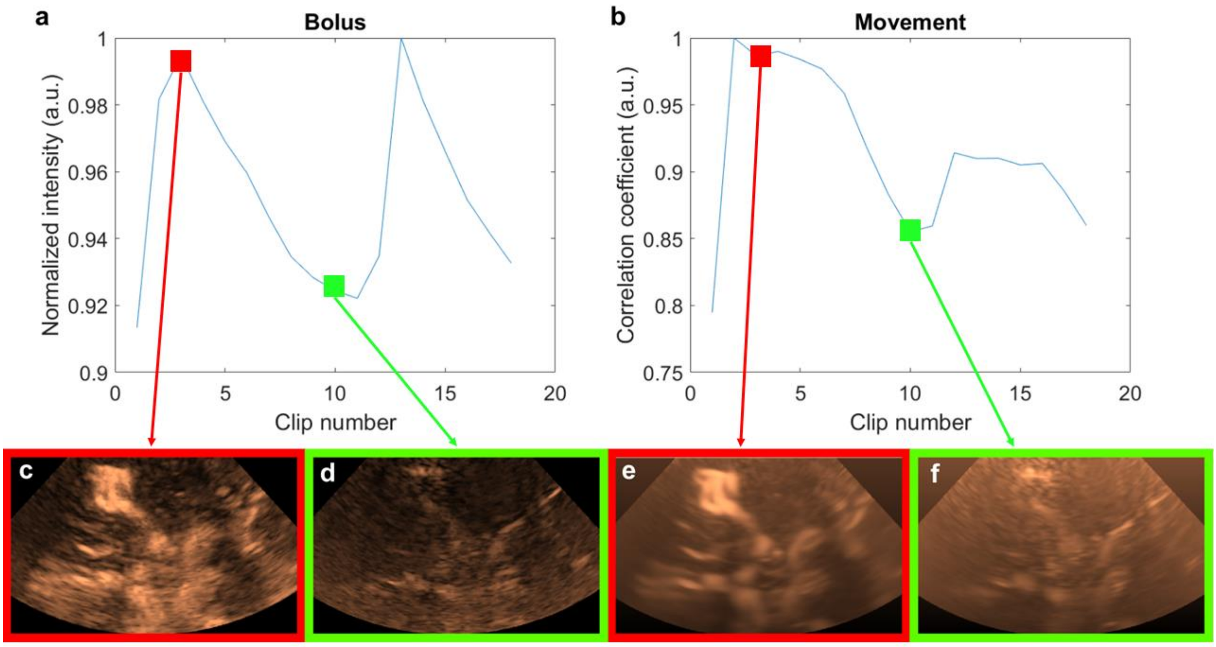


***Supplementary Figure 1. Estimation of MB concentration and probe movements in patient 1.*** *a) Concentration study: value of mean intensity normalized as a function of clip number, b) Movement study: correlation coefficient between the power Doppler of clip 2 and the power Doppler of the other clips, c) Peak of the first bolus (image from clip 2), d) End of first bolus (image from clip 10), e) Power Doppler of clip 2, f) Power Doppler of clip 10: a slight rightward drift can be observed.*

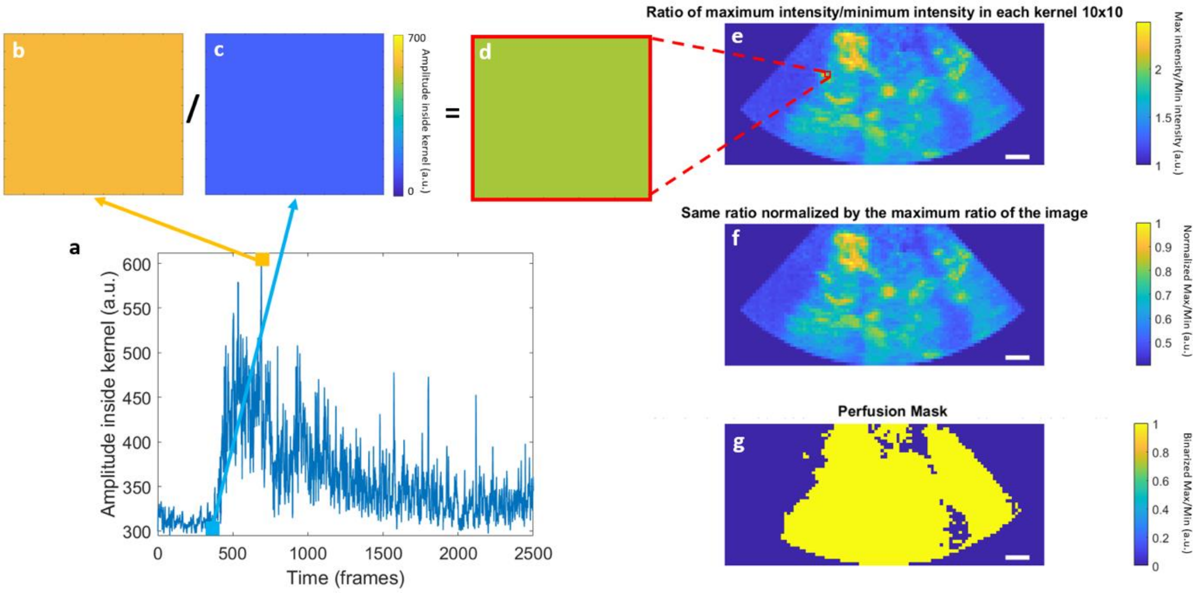


***Supplementary Figure 2. Creation of a perfusion mask to avoid localizing noise.*** *a) Perfusion curve (intensity vs. time) within the 10x10-pixel kernel zoomed in e), b) Maximum intensity of this 10x10-pixel kernel, c) Minimum intensity of this 10x10-pixel kernel, d) Result of dividing this kernel (maximum over minimum) zoomed in e), e) Maximum temporal intensity divided by the minimum temporal intensity in each of the 10x10 pixel kernels. The kernel shown in b, c, and d is framed in red, f) the Same ratio normalized by the maximum value, in the whole image, of this ratio, g) the Perfusion mask corresponding to this normalized ratio and binarized with a fixed threshold (i.e. the median of the normalized ratio of the image presented in f). 1cm scales.*

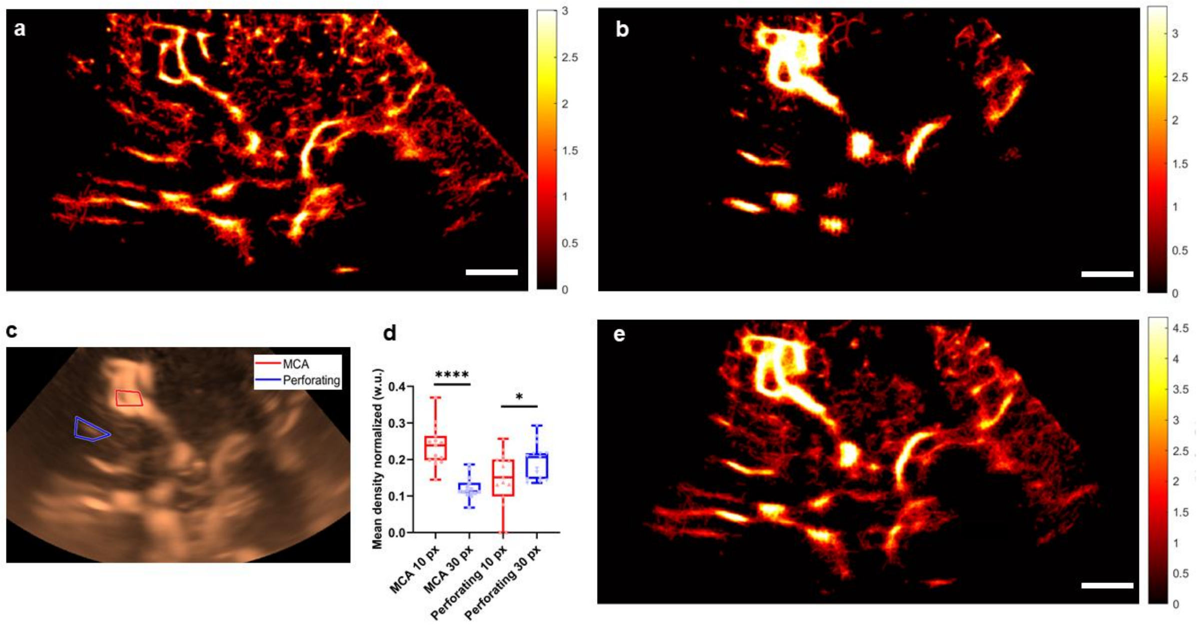


***Supplementary Figure 3. Influence of the size of the PSF on ULM density maps: the necessity of the double localization step. Study of 10 pixels (2 λ) PSF and 30 pixels (5 λ) PSF locations in the 15 control patients.*** *a) Vascular density mapping using the 10 pixels PSF criterion in patient n°1. Colormap in arbitrary units (a.u.), scale 1cm, b) Same for 30 pixels PSF locations, c) Segmentation of MCA and perforating on patient n°1, d) Comparison of intensity in each of the regions (MCA and perforating) on both 10 and 30 pixels PSF mapping in the 15 control patients. Student's t-test to compare MCA and perforating, e) Vascular density mapping performed by merging the two fusions of the two locations in patient n°1.*

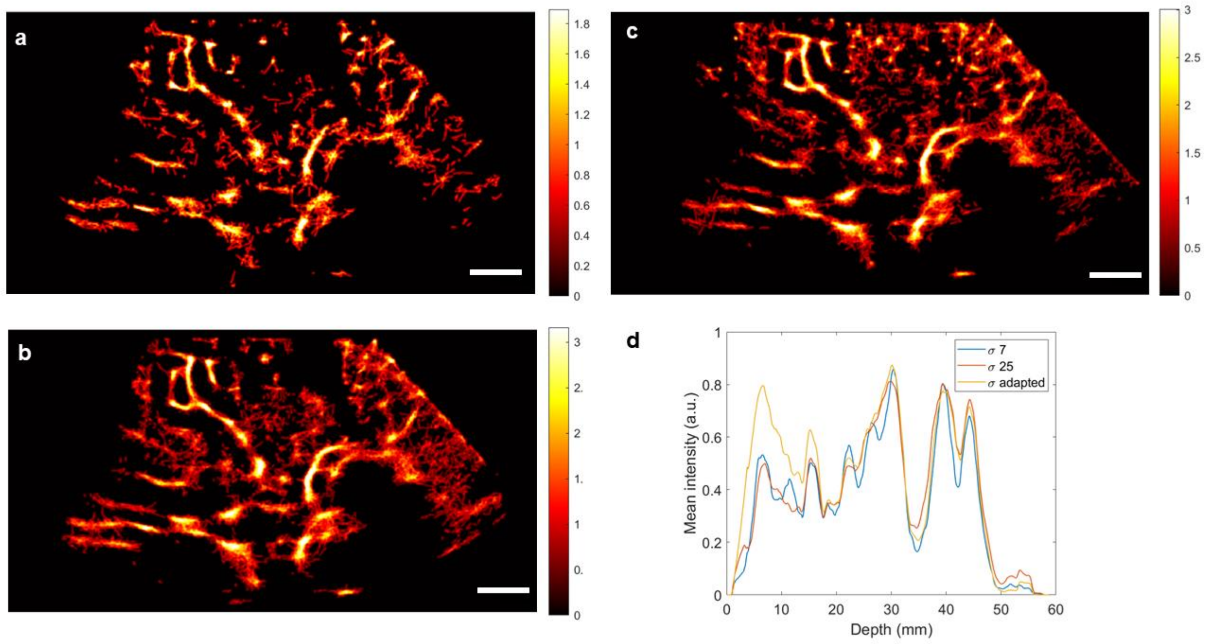


***Supplementary Figure 4. Influence of the σ of the PSF on ULM density maps: the necessity of the adapted σ as a function of depth.*** *a) For a PSF size set at 30 pixels (isolated), the standard deviation of the Gaussian PSF σ set at 7 seems to consider microbubbles too isolated, and therefore tends to create holes in the mappings, b) for the same PSF size using a σ of 25 pixels enhances vessels in-depth, c) for the same PSF size, the σ adapted in-depth seems to enhance low and high perforators, which is confirmed by the intensity analysis of density maps as a function of depth d). Colormap in arbitrary units (a.u.), scale 1cm.*

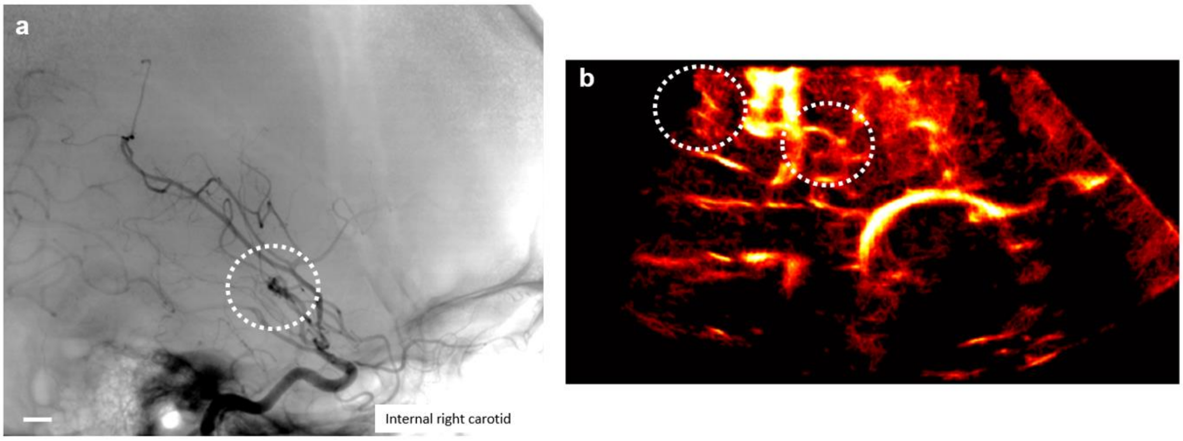


***Supplementary Figure 5. Comparison between arteriography and ULM on Moya Moya patient 4.*** *a) Time capture of arteriographic acquisition on the right MCA side. Sagittal plane, 1cm scale. b) ULM density map of the same Moya Moya patient. White dotted circles indicate the presence of the Moya Moya supply network in the two imaging modalities.*

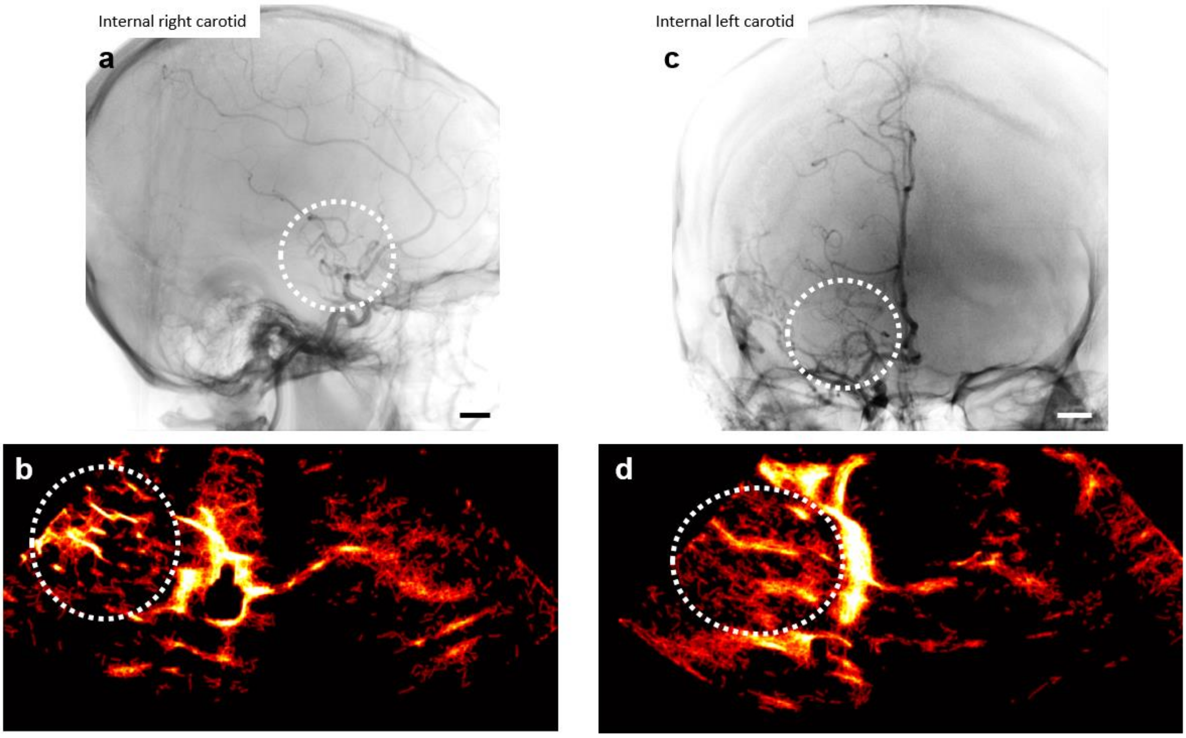


***Supplementary Figure 6. Comparison between arteriography and ULM on Moya Moya patient 7.*** *a) Time capture of arteriographic acquisition on the right MCA side. Sagittal plane, 1cm scale. b) ULM density map of the same Moya Moya patient. White dotted circles indicate the presence of the Moya Moya supply network in the two imaging modalities. c-d) Same patients, coronal plane, on the control left MCA side (unilateral Moya Moya).*
